# The cost of building an environmental surveillance system for typhoid

**DOI:** 10.1101/2021.07.12.21260395

**Authors:** Brittany L. Hagedorn, Nicolette A. Zhou, Christine S. Fagnant-Sperati, Jeffry H. Shirai, Jillian Gauld, Yuke Wang, David S. Boyle, John Scott Meschke

**Affiliations:** Bill & Melinda Gates Foundation, Institute for Disease Modeling; University of Washington, Environmental and Occupational Health; Emory University; PATH

## Abstract

**Introduction:** The typhoid conjugate vaccine is a safe and effective method for preventing Salmonella enterica serovar Typhi (typhoid) and the WHO’s guidance supports its use in locations with ongoing transmission. However, many countries lack a robust clinical surveillance system, making it challenging to determine where to use the vaccine. Environmental surveillance is an alternative approach to identify ongoing transmission, but the cost to implement such a strategy is previously unknown.

**Methods:** We estimated the cost of an environmental surveillance program for thirteen potential protocols. Unit costs were obtained from research labs involved in protocol development and equipment information was obtained from manufacturers and expert opinion. The cost model includes thirteen components and twenty-seven pieces of equipment. We used Monte Carlo simulations to project total costs.

**Results:** Total costs per sample including setup, overhead, and operational costs, range from $357–794 at a scale of 25 sites to $116–532 at 125 sites. Operational costs (ongoing expenditures) range from $218–584 per sample (25 sites) to $74–421 (125 sites). Eleven of the thirteen protocols have operational costs below $200 at this higher scale. Protocols with high up-front equipment costs benefit more from scale efficiencies. Sensitivity analyses show that laboratory labor, process efficiency, and the cost of consumables are the primary drivers of cost.

**Conclusion:** At scale, environmental surveillance may be affordable, depending on the protocol chosen and the geographic context. Costs will need to be considered along with protocol sensitivity. Opportunities to leverage existing infrastructure and multi-disease platforms may be necessary to further reduce costs.

**Summary Box:** *What is already known?:* Environmental surveillance has been used for polio transmission surveillance to target vaccination campaigns to prevent outbreaks. Similarly, methods for typhoid environmental surveillance are being developed, and could be used to support vaccine introduction decisions, if they are accurate and affordable.

*What are the new findings?:* Across the scenarios examined, operational costs are between $74–584 per sample depending on the scale and protocol selected. Operational costs are 67–79% of total annualized costs across the same permutations.

*What do the new findings imply?:* Policymakers can use these estimates and understanding of the efficiency of scale benefits in order to design a surveillance system that balances total cost with surveillance sensitivity and geographic coverage.

## Introduction

In March 2018, the World Health Organization (WHO) issued guidance^1^ supporting use of the recently developed typhoid conjugate vaccine (TCV) in populations at risk for typhoid fever. The TCV was pre-qualified by the WHO in December, making it available through Gavi, the Vaccine Alliance (Gavi) to qualifying low-income countries. The introductions began in 2019, with Pakistan being the first country in the world to include it in their routine immunization schedule^2^.

Currently, the majority of Gavi-qualified countries have few, if any, sites that conduct routine clinical surveillance for typhoid fever, resulting in estimates of global burden that are based on very few reporting sites^3,4^. With many competing health priorities, generating evidence of the true burden of disease is critical for countries trying to decide when and if to introduce TCV. The information available to assess sub-national burden is even less, making it difficult to consider a targeted vaccination strategy and to assess the impact of a TCV roll-out. Because of the limited feasibility and high cost of long-term expansion of clinical surveillance in these settings, environmental surveillance (ES) is being considered as a suitable alternative for reliable and continued monitoring, although protocols are still in development and used for research purposes^5–7^.

Previous work has examined the total cost of the global polio laboratory network (including both environmental and case surveillance) and suggested that true total costs were higher than budgeted^8^, but they are also subject to economies of scale^9^. Given this, cost estimates for a future typhoid ES system must consider costs of implementation not for a single sampling site, but when expanded to cover large geographic regions and higher volumes. Additionally, costing studies from other parts of the healthcare system in low- and middle-income countries (LMIC) have shown that the delivery and overhead costs may account for 31-87% of site-level costs^10^, highlighting the need to factor them into estimates of future surveillance costs.

Due to the ongoing nature of the development and field testing of ES protocols for typhoid, there remains a substantial amount of variation in sample processing and laboratory methods, which needs to be accounted for. The survey of labs developing typhoid environmental surveillance protocols showed that protocols were highly variable with regard to sample collection method, concentration, enrichment steps, and analysis methods; the thirteen identified protocols from seven laboratories are summarized in Table 1.

**Table 1.**
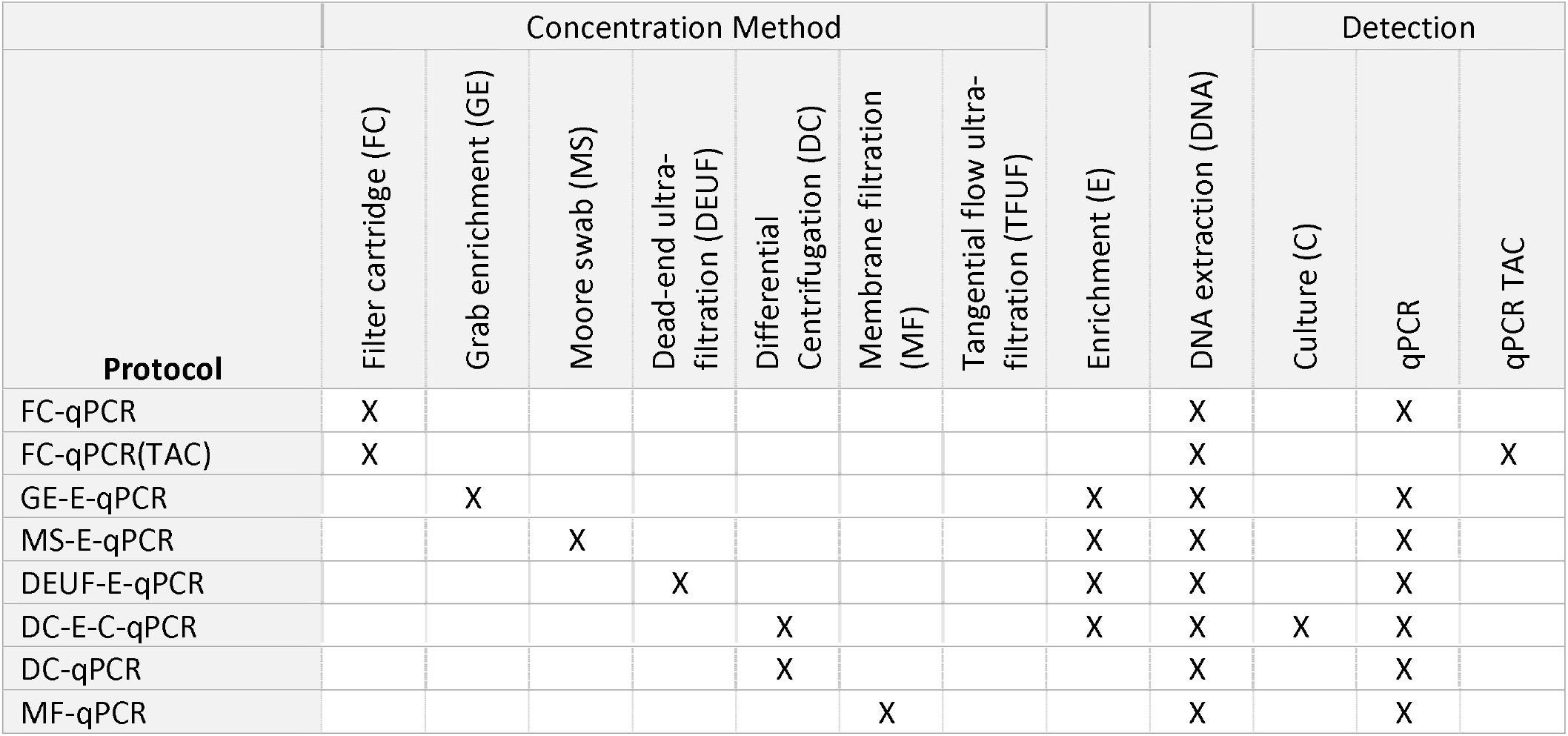
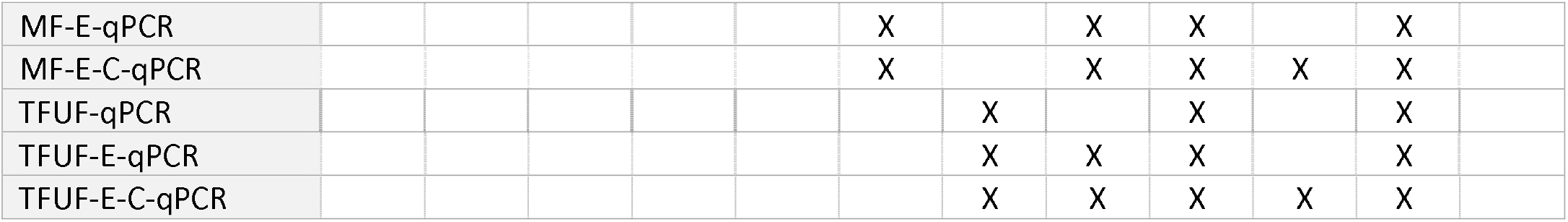
Protocols (named in the rows) are composed of multiple methods (names in columns). X intersections represent methods that are included in each protocol and are relevant for costing purposes. Each method (in the columns) requires different resources, which impact the cost of sample collection and processing. FC = filter cartridge. GE = grab enrichment. MF = membrane filtration. MS = Moore swab. TFUF = tangential flow ultrafiltration.

The methods summarized in Table 1 may have differing sensitivity and specificity, cost, and operational scalability, which would all impact how successfully they can be implemented in a routine monitoring system. This is valuable to inform accurate budgeting, but more importantly, it is needed in order to guide realistic target-setting for ‘acceptable’ costs per sample as part of target product profiles (TPPs) and for informing discussions on how much of the total cost an LMIC will be able to fund themselves, and thus how much donor support may be needed.

This paper explores the drivers of costs for different protocols and quantifies the differences in both per-sample cost and the efficiency of scaling up to higher numbers of samples. By estimating the total costs across multiple protocols, any comparisons between them will then be on equal footing, since the estimates will all include the major cost drivers and have comparable assumptions, thus informing trade-off discussions and future research and development strategies.

## Methods: Cost Model

The cost of a full-scale typhoid ES laboratory system is currently unknown, since it has not yet been built, but there is a need to understand the magnitude of the total costs in order to begin planning for funding and implementation. Here, we made the first estimate of the total and per-sample costs for thirteen protocols that are currently in development or use (Table 1 and Supplement 1). We estimated the full cost to run a laboratory dedicated only to typhoid ES, which resulted in conservative (***i*.*e***., most expensive) estimates. We took a programmatic perspective and included financial costs only. All values are reported in 2019 U.S. dollars.

### Data collection and assumptions

We made use of unit cost surveys (Supplement 3) conducted by the Environmental and Occupational Health Microbiology Laboratory at the University of Washington, as part of their evaluation of typhoid ES protocols, and when data were not available, estimates were informed by expert opinion. Survey respondents included seven labs conducting a total of thirteen protocols in Bangladesh, India, Kenya, Malawi, Nepal, and Pakistan. Costs were reported in the survey in US dollars based on the laboratories’ real expenditures. Because survey responses included details on specific lab research budgets, we aggregated survey results so that individual labs cannot be identified. The first quartile, median, and third quartile were used to parameterize the uncertainty in the distribution of unit cost values.

The total cost model was a generalized exploration of total costs and unit costs at scale. It did not consider localized effects specific to countries, such as import duties on equipment, fuel rate variations, or tax rates on various components of the input costs.

The cost model included components for labor, equipment, maintenance, consumables, depreciation, and overhead. Each of these had multiple inputs as described below and were calculated using an ingredients-based approach. Surveys reported per-sample unit and consumables costs (***e*.*g***., reagents), required equipment, labor time per sample, and samples collected per day per team. Expert opinion was provided by personnel adept in conducting the field and laboratory methods, and informed the list of required equipment, equipment capacities, labor time per sample, laboratory batch size, technicians per team, and samples collected per day per team. Available commercial pricing in the United States informed consumables and equipment costs where there were gaps in the survey data. A comprehensive listing of all equipment requirements (Table S1.1), unit costs (Tables S1.3 and S1.5), equipment capacities and lifespans

(Table S1.2), labor time per sample (Table S1.4), laboratory batch size (Table S1.4), technicians per team (Table S1.4), and samples collected per day per team (Table S1.4) is in the supplement.

For the purposes of this paper, the term ‘method’ refers to a single step in the protocol, such as DNA extraction, and the term ‘protocol’ refers to the combination of methods that make up the full processing of a sample.

### Model structure

We calculated the year-one investment required for capital expenses (*e*.*g*., new equipment purchases) and the recurrent operational costs separately; these sum to the total cost.

The model components are shown in Figure 1 and the formulaic structure of the model is detailed in Supplement 4. For each cost component, whenever the number of samples being processed exceeded capacity, we incremented the number of processing units until it was adequate. For example, if a particular piece of equipment had a weekly capacity of 100 samples, then to process 101 samples, we assumed the purchase of a second piece of equipment and incremented the cost. The same was true for the number of collection teams and laboratory FTEs; when the number of samples exceeded the capacity of the staff, there was incremental cost to bring in additional staff to meet these needs.

**Figure 1.**
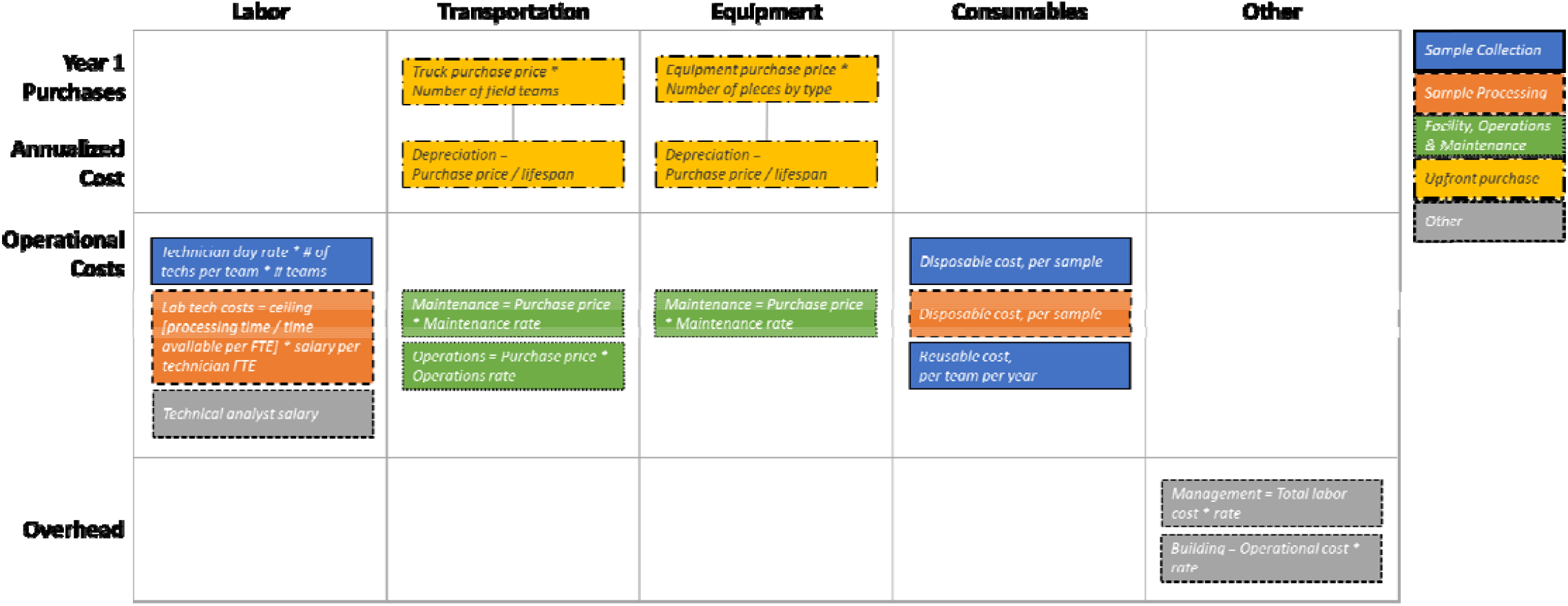
Cost model components, categorized by type and color-coded by function. Detailed descriptions and formulas can be found in Supplement 4. Operational costs include labor, truck operations and maintenance, equipment maintenance, and consumables.

For this modeling exercise, we assumed bi-weekly sampling at each collection site (*i*.*e*., 26 samples per site per year), that staff typically work 46 weeks per year (*i*.*e*., four weeks of vacation and two weeks of holidays), 5 days per week, and that laboratory staff are productive during 80% of their working hours (*i*.*e*., the remainder is spent on non-productivity tasks such as trainings, planned maintenance, supply management, etc.). We assumed that sample collection staff are flexible and could be paid on a daily-rate basis. We assumed that laboratory staff were hired full time, regardless of their utilization level. Management and building overhead costs were estimated with a traditional approach, multiplying the appropriate percentage to other component costs.

The model was built in R Studio version 1.3.959^11^ and using the R program version 3.6.3^12^. Code is available on GitHub upon request.

### Model application

The cost model simulated 1,000 trials for each protocol of varying collection and laboratory sample processing methods. Each unit cost, time duration, and percentage value were sampled for each trial as part of a Monte Carlo simulation from a truncated normal distribution. For each distribution, we used the survey-reported variation in values; where there were too few data points available, we assumed a standard deviation of twenty percent. For example, for each of the 27 potential pieces of equipment there is an equipment-piece-specific maintenance rate pulled from the appropriate distribution, each with the same expected value but varying because of the stochastic draws; these were multiplied by the base purchase prices (which were also sampled), respectively.

To calculate the total cost per sample processed and assess the impact of scale on the per-sample cost, we assumed a constant use of a single central laboratory facility, but varied the number of samples collected, assuming bi-weekly collection (***i*.*e***., 26 samples per site per year) and varying the number of sampling sites from 25 to 125, to represent geographic coverage from a small region to a country-wide strategy. Increases in the number of sites impacted the number of collection teams and trucks required to reach these sites; if additional equipment or laboratory staff were required to process the increased volume of samples, those were also included in the cost-per sample calculations. These requirements are summarized in Supplement 2.

Total annualized costs and operational costs are the two primary results. Operational costs included labor, truck operations and maintenance, equipment maintenance, and consumables (including both reusables and disposables); these are expenses that would need to be budgeted on an ongoing basis. Total annualized costs also included the purchase price for trucks and equipment in the form of depreciation, plus management and building overhead expenses.

Given the uncertainty inherent in the ongoing development of an ES protocol for routine use, we also conducted a sensitivity analysis to assess the impact of factors on either process or unit costs. This sensitivity analysis examined the impact of a 20% change in values for each of the following factors: samples collected per team per day, equipment capacity maximums, laboratory processing time per batch, depreciation lifespan for equipment and trucks, management and building overhead cost rates, collection labor pay rates, equipment and truck purchase costs, equipment and truck maintenance and operational costs, and consumables costs per team and sample.

## Results

### Cost per sample by protocol

The modeled costs for each protocol are shown in Figure 2 and the values are quantified in Supplement 2. Annualized total costs represent the fully loaded expense of setting up, operating and maintaining the surveillance system. Operational costs are a subset of those, representing the ongoing direct expense of running the system, thus they are always lower than the annualized values. The expected average cost per sample is shown by the size of the bar and varies by protocol, with some being much higher than others. However, these results are for cost only and do not account for the sensitivity of the protocol, which could result in expensive protocols being more cost-effective.

**Figure 2.**
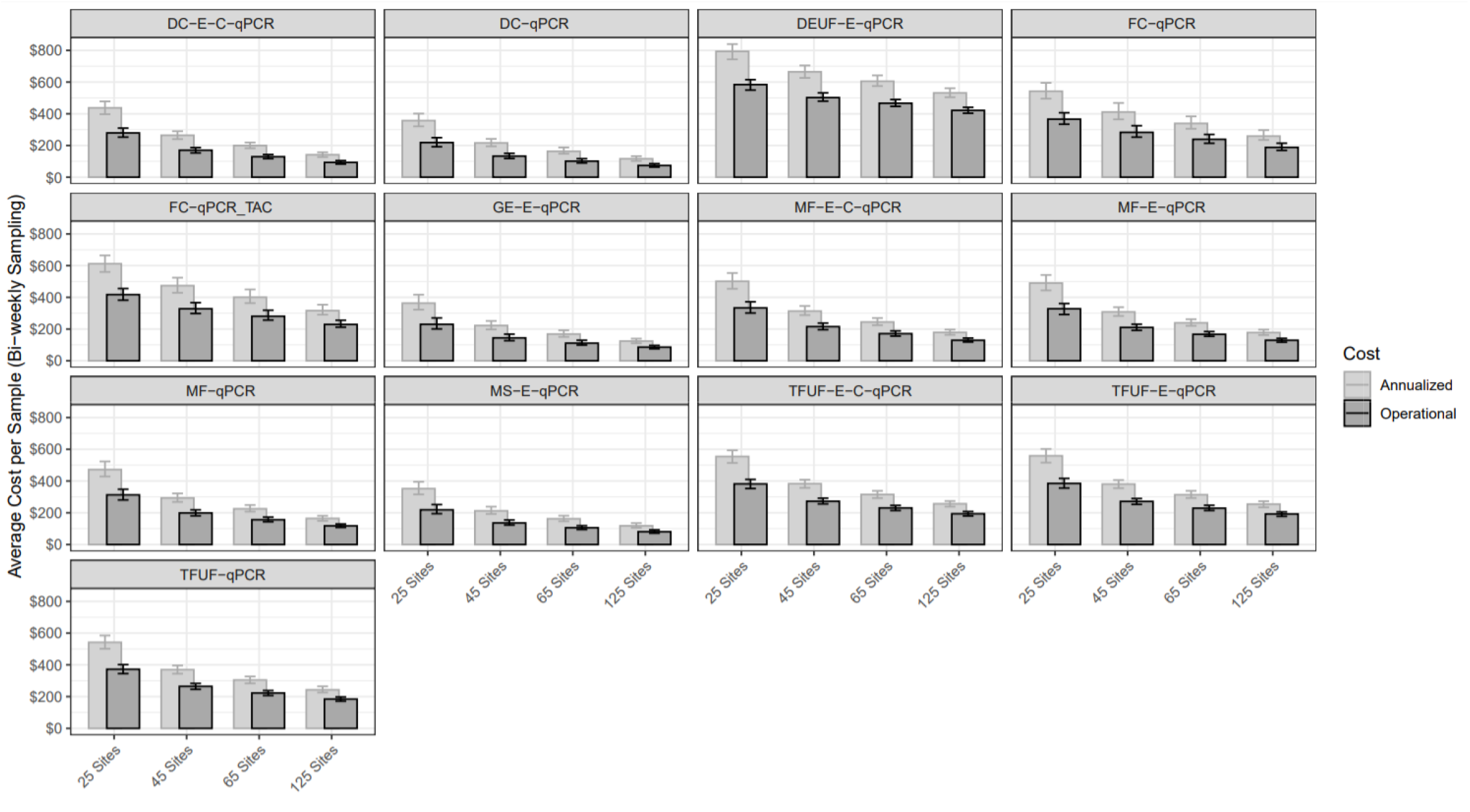
Total annualized cost (inclusive of all system costs) and operational costs for an example number of twenty-five, forty-five, sixty-five, and a hundred and twenty-five sites, sampled bi-weekly. All values in 2019 US dollars. Error bars represent the 25 ^th^ and 75^th^ percentile of simulated results. DC = differential centrifugation. DEUF = dead end ultrafiltration. FC = filter cartridge. GE = grab enrichment. MF = membrane filtration. MS = Moore swab. TFUF = tangential flow ultrafiltration.

Since the magnitude of a potential surveillance system is unknown, we calculate the per-sample cost at varying levels of sample numbers and assess the impact of economies of scale on the unit cost per sample. Most of the cost savings on a per-sample basis happen with relatively modest increases in sample numbers. It is important to note that the relative cost of protocols does not remain constant with scale; for example, FC-qPCR benefits from scale faster than DEUF-E-qPCR, while the MS-E-qPCR protocol is consistently a lower-cost protocol per sample.

### Total annualized cost and operational cost per sample for each protocol are reported in Supplement 2

The total cost is the aggregation of six components: labor, equipment, trucks, consumables, management overhead, and the laboratory building (Figure 1). Each of these varies at a different rate and thus benefit from economies of scale to varying degrees. For example, consumables consistently rise as a fraction of the total cost as scale increases bec use the unit costs per sample are essentially fixed. In comparison, the fraction of the total cost that is spent on equipment and labor decreases with scale as large up-front expenses (*e*.*g*., equipment that has a high capacity) are spread across incremental samples. (See Figure 4.)

**Figure 4.**
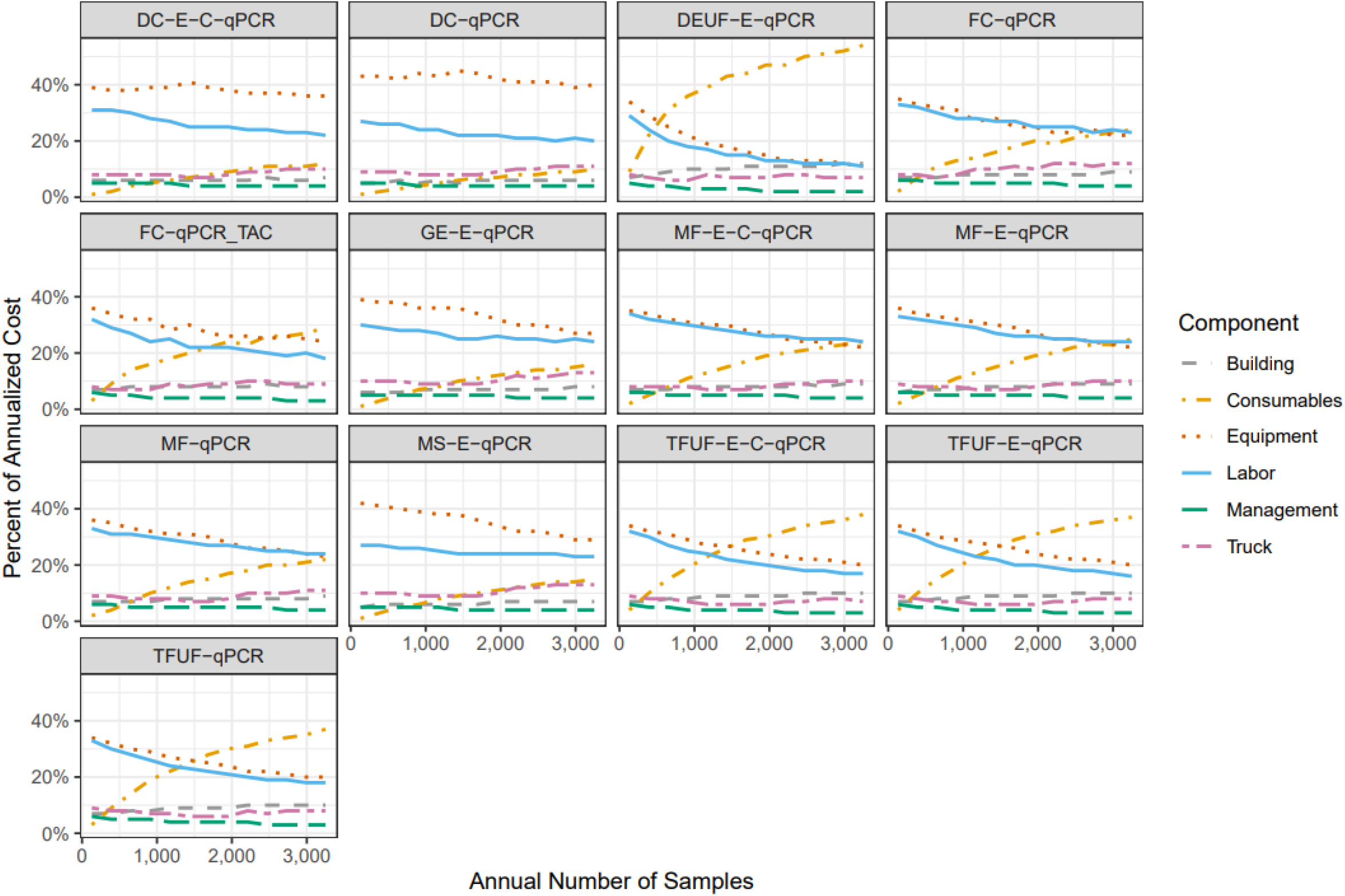
Breakdown of annualized costs as a percentage of the total for each protocol. Values are the median of the percentage calculations from 1,000 simulation trials, so the component categories may not sum to 100%. Non-smooth trajectories are indicative of step-changes in cost, for example the purchase of an additional piece of equipment or the need to hire a staff member. Assumes bi-weekly sampling and that the increase in number of samples is due to incremental sampling sites. DC = differential centrifugation. DEUF = dead end ultrafiltration. FC = filter cartridge. GE = grab enrichment. MF = membrane filtration. MS = Moore swab. TFUF = tangential flow ultrafiltration.

For protocols relying on filter cartridges, dead-end ultrafiltration, and tangential flow ultrafiltration, the cost of consumables becomes the largest component of total cost once a laboratory reaches scale. Other protocols, such as those relying on differential centrifugation, have high upfront equipment investment requirements that continue to be a large component of total cost even at scale. For all protocols, labor is consistently a top-two contributor to costs.

### Sensitivity analysis

Since there is still work in progress to finalize the protocols and implement routine ES, we conducted a sensitivity analysis to identify which of the key cost components have the most impact on the annualized cost per sample. We applied a twenty percent adjustment to each of the relevant values and the results are shown in Figure 5. The vertical axis categories are ordered according to their aggregate impact across all of the protocols. Two of the three most impactful values are related to the laboratory staff: labor costs (pay rates) and lab processes (how much time a single batch takes to process, which impacts the number of staff needed). Consumable prices also have substantial impact on uncertainty. The equipment- and truck-related costs fall at the bottom of the list, in decreasing order starting from depreciation lifespan, purchase cost, maintenance and operations rates, and equipment capacity.

**Figure 5.**
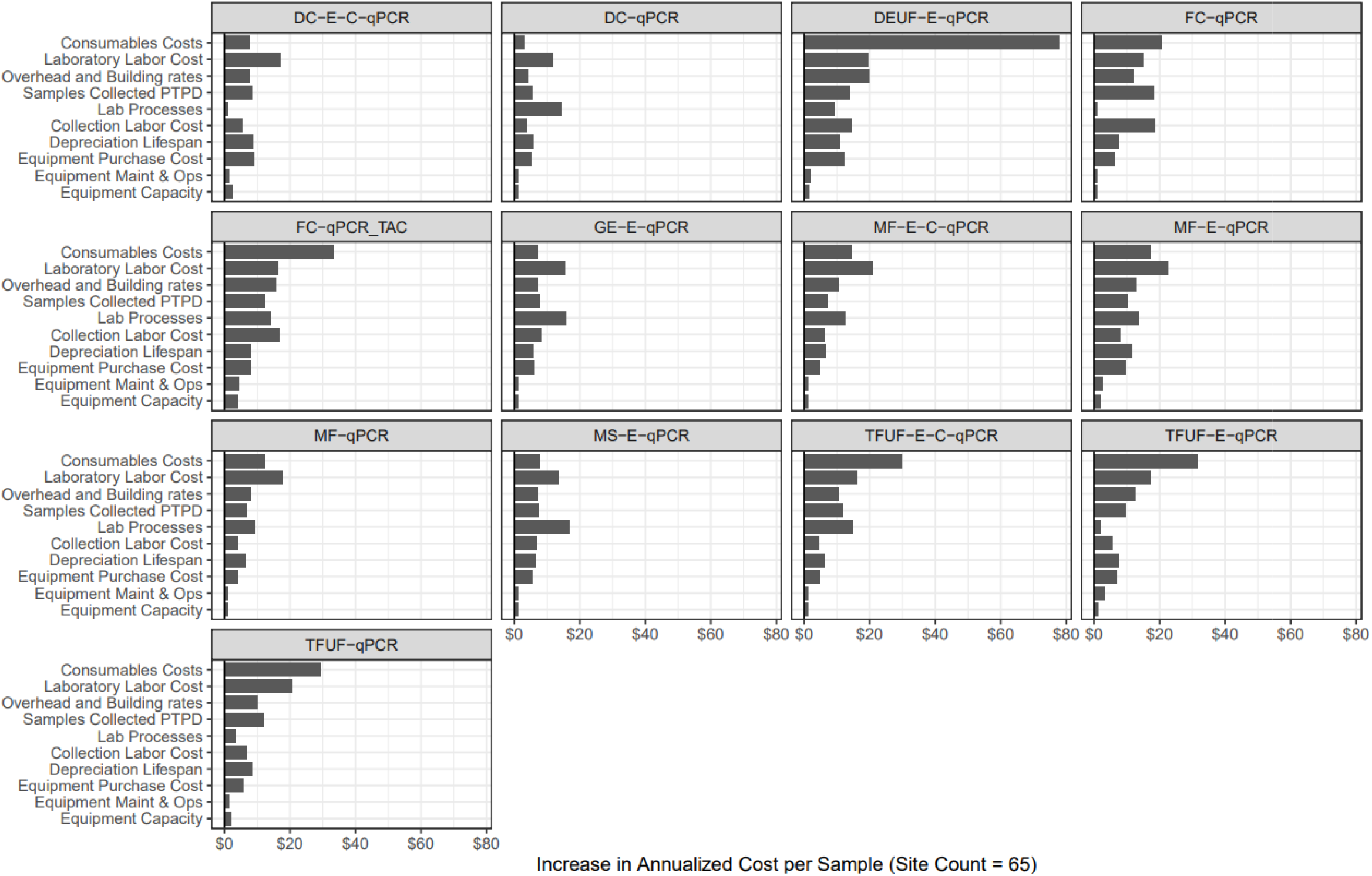
Sensitivity analysis reflecting the impact of a 20% change in the relevant baseline value and its impact on the annualized cost per sample. For example, laboratory labor cost multiplies the salary values by 1.2 to increase the total; depreciation lifespan multiplies the number of years by 0.8, which thus increases costs due to faster replacement. All values in 2019 US dollars. Values represent an example scenario with sixty-five sites and bi-weekly sampling (*i*.*e*. 26 samples per year per site). Increases are calculated as the difference between the mean baseline and 20% adjusted scenarios. DEUF = dead end ultrafiltration. FC = filter cartridge. GE = grab enrichment. MF = membrane filtration MS = Moore swab. TFUF = tangential flow ultrafiltration.

## Discussion

The thirteen protocols compared in this study utilize different types of equipment, varied levels of labor intensity, and consumables costs. As a result, the cost per sample can vary widely. At a level of 25 sites, median operational cost per sample ranges from $218 to $584. This decreases with scale; at 125 sites, the range of median operational cost per sample is $74 to $421, with eleven of the thirteen protocols coming in below $200.

In addition to differences between protocols, there is also parameter uncertainty, which we addressed by using a Monte Carlo simulation, with distributions based on the range of unit costs reported by participating labs. These ranges were found to be quite substantial in some cases and the effect on total costs was more pronounced at lower sample volumes.

Economies of scale were a primary driver of the cost per sample, both operational-only and total costs. This was true across all protocols, as up-front costs of labor and equipment are spread over larger numbers of samples. Substantial cost-per-sample reductions were seen up to ∼1000 samples per year; after that they continued but at a slower pace due to having achieved most of the efficiencies from large capacity equipment.

However, it is important to note that not all protocols scale in the same way, with some benefiting more than others depending on their mix of semi-fixed labor and equipment costs versus less scalable consumables and collection costs. This has implications for funding mechanisms. In many LMICs, surveillance programs are funded in part through donor support. If countries are eventually expected to pay for the ongoing operations of their surveillance programs, their preferences may be toward protocols that have high up-front investments but lower ongoing costs, pushing them away from methods with high consumables (such as those using DEUF or TFUF) or laboratory labor costs, for example.

There may also be additional cost savings possible through other means. For example, group purchasing options for laboratory supplies and equipment could result in substantial savings. Bulk procurement at discount rates for equipment and reagents is already done by UNICEF’s Supply Division, the US Centers for Disease Control and Prevention (CDC), the President’s Emergency Plan For AIDS Relief (PEPFAR), and the United States Agency for International Development (USAID); a similar strategy could be employed for ES-related purchasing.

Scale was the largest driver of the cost-per-sample, but we also found via the sensitivity analysis that protocols were particularly sensitive to the cost of laboratory labor, both pay rates and the time to process a sample. Salaries are variable by country, so this implies that the cost of an ES program is substantially dependent on the labor market. This is may reduce total cost if the surveillance system is being used in a low-income country where salaries are generally lower, but it does mean that the cost estimates presented here are sensitive to context.

Although the cost of equipment has implications for economies of scale, these costs were less impactful in the sensitivity analysis and the most important factors were the lifespan and the purchase price. These are the two variables required to calculate annual depreciation, which is the largest cost related to equipment and trucks overall. The lifespans used were based on depreciation standards, but that may not reflect practical durability, especially in LMIC contexts if there are less well-controlled, harsher operating environments, or limited maintenance^14^. On the other hand, resource-limited environments may also be forced to utilize laboratory equipment and/or supplies for longer than the depreciation estimates, which too can impact cost projections.

As protocols are further refined, these estimates will need to be updated with more accurate input values, with a priority on the parameters highlighted through the sensitivity analysis. Equipment is 30-40% of annualized total costs at low volumes and since equipment is a large part of initial investments, details about which manufacturer is approved for use will be critical. For global surveillance programs, tests are often developed and validated for use on certain platforms and thus the manufacturers’ decisions about how to set prices (including technical support and maintenance) will have substantial impact and need to be incorporated once they are finalized. Additional refinements for local labor costs, equipment importation expenses, local distributor procurement mark-ups, and quality assurance programs would also improve accuracy.

The operational model will also impact effectiveness, as there is the potential for sample failure due to prolonged time between sample collection and laboratory receipt with the potential for cold-chain failure during shipment. This leads to trade-offs between a laboratory system design that relies on smaller labs with less opportunity to benefit from cost efficiencies, as compared with centralized labs that are lower cost but may introduce opportunities for sample failure due to longer shipment times from distributed sampling locations.

Of course, cost is not the only factor in a decision about which protocol to adopt. The approach that is ultimately selected must be 1) feasible in the local context (***e*.*g***., trained staff and key reagents are available), 2) reliable despite possible logistical and laboratory delays (***e*.*g***., shipping from remote locations), and 3) both sensitive and specific enough to meet surveillance needs. Thus, once the protocols have been laboratory- and field-validated, the costs will need to be balanced with effectiveness. Future work should use measures such as the lower limit of detection or the number of samples (or sites) required to meet a certain use case.

## Conclusion

This study lays out the framework to investigate drivers of costs for environmental surveillance across a range of methodological approaches and protocols. Although variability exists between protocols’ overall costs, there is a general observation that the economies of scale for environmental surveillance may be significant. With awareness of this dynamic, future policymakers can use this information to design a surveillance and laboratory system that optimizes existing resources, and to consider how broadening surveillance may scale from a cost perspective.

## Data Availability

All data is available within the manuscript and supplements. Input assumptions and cost data from the survey of laboratories is summarized and available in the supplementary materials. Detailed results are also summarized in the supplementary materials and visualized within the main document.
Inquiries for additional details can be made to the corresponding author ORCID iD 0000-0002-1489-7168.

## Acknowledgements

We would like to thank Supriya Kumar for her contributions to understanding the need for this study and what information would be most useful. We would also like to thank all of the members of the Typhoid environmental surveillance working group for their invaluable contributions and willingness to contribute unit cost and protocol information for use in this study.

## Supplement 1: Cost Model Input Parameters

**Table S1.1.**
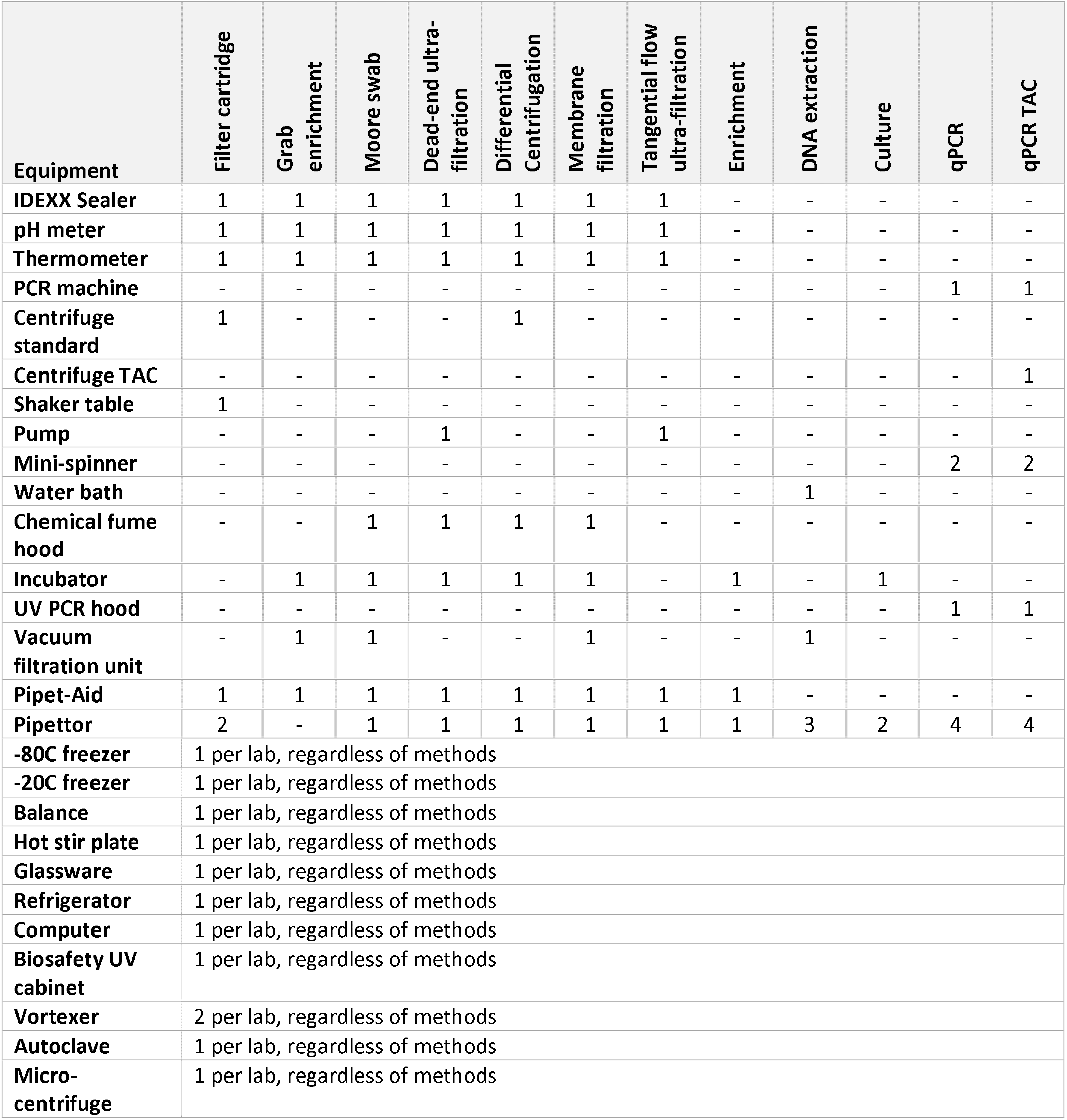
Method-based laboratory equipment requirements. The number indicates how many of each piece of equipment is needed for a given aspect of the environmental surveillance method protocol. Equipment needs by method were obtained through surveys of participating laboratories.

**Table S1.2.** Capacity of laboratory equipment. Daily capacity represents the maximum number of samples that could be processed by that piece of equipment in a single day. Lifespan (samples) represents the maximum number of samples that could be processed by that piece of equipment before it needs to be replaced. Lifespan (years) represents the maximum number of years that a piece of equipment could be in place before it would need to be replaced; duration is consistent with the US IRS guidance on depreciation. Values were obtained through surveys of participating laboratories and online research for each piece of equipment.

| Equipment | Daily capacity (samples) | Daily capacity (teams) | Lifespan (samples) | Lifespan (years) |
| --- | --- | --- | --- | --- |
| IDEXX Sealer | - | - | 5,000 | - |
| pH meter | - | - | 1,000 | - |
| Thermometer | - | - | 5,000 | - |
| PCR machine | 300 | - | - | 5 |
| Centrifuge standard | 50 | - | - | 5 |
| Centrifuge TAC | 50 | - | - | 5 |
| Shaker table | 50 | - | - | 5 |
| Pump | 10 | - | - | 5 |
| Mini-spinner | 300 | - | - | 5 |
| Water bath | 500 | - | - | 5 |
| Chemical fume hood | 1,000 | - | - | 5 |
| Incubator | 100 | - | - | 5 |
| UV PCR hood | 600 | - | - | 5 |
| Vacuum filtration unit | 500 | - | - | 5 |
| Pipet-Aid | 600 | - | - | 5 |
| Pipettor | 600 | - | - | 5 |
| -80C freezer | 20,000 | - | - | 5 |
| -20C freezer | 15,000 | - | - | 5 |
| Balance | 1,000 | - | - | 5 |
| Hot stir plate | 1,000 | - | - | 5 |
| Glassware | 1,000 | - | - | 5 |
| Refrigerator | 300 | - | - | 5 |
| Computer | 10,000 | - | - | 5 |
| Biosafety UV cabinet | 600 | - | - | 5 |
| Vortexer | 600 | - | - | 5 |
| Autoclave | 200 | - | - | 5 |
| Micro-centrifuge | 100 | - | - | 5 |
| Truck | - | 1 | - | 7 |

**Table S1.3.** Unit costs for consumables. Disposable costs include all supplies and wastage related to the given method, as reported via surveys of participating laboratories. All values in US dollars, 2019.

| Value |  | Filter cartridge | Grab enrichment | Moore swab | Dead-end ultra-filtration | Differential Centrifugation | Membrane filtration | Tangential flow ultra-filtration | Enrichment | DNA extraction | Culture | qPCR | qPCR TAC |
| --- | --- | --- | --- | --- | --- | --- | --- | --- | --- | --- | --- | --- | --- |
| Disposables | 25<sup>th</sup> | 60.00 | 1.50 | 0.80 | 283.50 | 0.80 | 27.00 | 51.50 | 2.50 | 1.80 | - | 2.00 | 45.00 |
|  | Median | 70.00 | 3.00 | 1.00 | 315.00 | 1.00 | 30.00 | 102.00 | 10.00 | 4.50 | - | 11.00 | 55.00 |
|  | 75<sup>th</sup> | 80.00 | 4.50 | 1.20 | 346.50 | 1.20 | 33.00 | 103.00 | 17.00 | 20.00 | - | 20.00 | 65.00 |
| Reusables, annual per team | 25<sup>th</sup> | 8.50 |  |  |  |  |  |  |  |  |  |  |  |
|  | Median | 17.00 |  |  |  |  |  |  |  |  |  |  |  |
|  | 75<sup>th</sup> | 117.00 |  |  |  |  |  |  |  |  |  |  |  |
| Reusables, daily per team | 25<sup>th</sup> | 1.50 | 1.50 | - | - | 1.50 | 9.00 | 51.50 | - | - | - | - | - |
|  | Median | 3.00 | 3.00 | - | - | 3.00 | 18.00 | 103.00 | - | - | - | - | - |
|  | 75<sup>th</sup> | 4.50 | 4.50 | - | - | 4.50 | 48.00 | 103.00 | - | - | - | - | - |

**Table S1.4.** Operational parameters used for estimating the number of labor-hours required to collect and process samples. Active lab staff time in hours. Blanks are spaces where the data are not applicable; these are not methods associated with collection. Values obtained via surveys of participating laboratories.

| Value |  | Filter cartridge | Grab enrichment | Moore swab | Dead-end ultra-filtration | Differential Centrifugation | Membrane filtration | Tangential flow ultra-filtration | Enrichment | DNA extraction | Culture | qPCR | qPCR TAC |
| --- | --- | --- | --- | --- | --- | --- | --- | --- | --- | --- | --- | --- | --- |
| Samples collected per team per day | Min | 1 | 2 | 2 | 1 | 2 | 2 | 2 | - | - | - | - | - |
|  | Mode | 2 | 5 | 5 | 2 | 5 | 5 | 5 | - | - | - | - | - |
|  | Max | 4 | 10 | 10 | 4 | 10 | 10 | 10 | - | - | - | - | - |
| Technicians per collection team |  | 3 | 2 | 2 | 2 | 2 | 2 | 2 | - | - | - | - | - |
| Laboratory batch size |  | 6 | 12 | 20 | 6 | 16 | 6 | 6 | 20 | 12 | 20 | 80 | 8 |
| Active lab staff time, per batch | Min | 1.5 | 0.5 | 0.3 | 0.5 | 1.0 | 0.5 | 1.5 | 0.3 | 1.0 | 0.5 | 1.0 | 1.0 |
|  | Mode | 2.0 | 1.0 | 0.5 | 1.0 | 1.5 | 2.0 | 2.0 | 0.5 | 2.0 | 1.0 | 2.0 | 2.0 |
|  | Max | 3.0 | 2.0 | 1.0 | 2.0 | 2.0 | 6.0 | 3.0 | 1.0 | 4.0 | 2.0 | 3.0 | 3.0 |

**Table S1.5.** Rates and values for the cost model.

| Parameter | Value | Reference |
| --- | --- | --- |
| Equipment maintenance rate | 4% | Assumed |
| Truck maintenance rate | 16% of depreciation | Calculated per <sup>13</sup> |
| Truck annual operations cost rate | 35% of depreciation | Calculated per <sup>13</sup> |
| Daily pay rate for technicians | \$3.50, \$7.00, \$40.00 | Survey of participating labs |

## Supplement 2: Total and Operational Costs per Sample

**Table S2.1.** Total and operational costs per sample at selected levels of scale and for each protocol considered. Annualized costs per sample represent the median, 25^th^, and 75^th^ percentile results from 1,000 simulation runs. All values are in 2019 US dollars.

| Protocol | Site Count,<br>Sampled Bi-Weekly | Annualized cost per sample<br>USD Median (25<sup>th</sup>, 75<sup>th</sup>) | Operational cost per sample<br>USD Median (25<sup>th</sup>, 75<sup>th</sup>) |
| --- | --- | --- | --- |
| DC-E-C-qPCR | 25 | 437 (397, 478) | 279 (253, 309) |
| DC-E-C-qPCR | 45 | 264 (240, 290) | 170 (153, 186) |
| DC-E-C-qPCR | 65 | 200 (182, 219) | 129 (117, 143) |
| DC-E-C-qPCR | 125 | 141 (127, 157) | 93 (83, 105) |
| DC-qPCR | 25 | 357 (321, 401) | 218 (192, 249) |
| DC-qPCR | 45 | 216 (195, 241) | 133 (118, 151) |
| DC-qPCR | 65 | 163 (148, 188) | 101 (90, 117) |
| DC-qPCR | 125 | 116 (102, 133) | 74 (64, 85) |
| DEUF-E-qPCR | 25 | 794 (744, 840) | 584 (549, 615) |
| DEUF-E-qPCR | 45 | 665 (626, 705) | 502 (480, 532) |
| DEUF-E-qPCR | 65 | 605 (575, 642) | 467 (447, 490) |
| DEUF-E-qPCR | 125 | 532 (504, 561) | 421 (403, 441) |
| FC-qPCR | 25 | 542 (495, 595) | 366 (334, 407) |
| FC-qPCR | 45 | 411 (365, 468) | 283 (253, 325) |
| FC-qPCR | 65 | 339 (305, 384) | 239 (214, 270) |
| FC-qPCR | 125 | 259 (235, 297) | 188 (169, 214) |
| FC-qPCR_TAC | 25 | 612 (560, 665) | 417 (382, 456) |
| FC-qPCR_TAC | 45 | 474 (429, 524) | 328 (298, 367) |
| FC-qPCR_TAC | 65 | 401 (363, 450) | 281 (257, 319) |
| FC-qPCR_TAC | 125 | 316 (290, 354) | 229 (212, 256) |
| GE-E-qPCR | 25 | 363 (322, 417) | 230 (201, 270) |
| GE-E-qPCR | 45 | 222 (198, 251) | 144 (127, 168) |
| GE-E-qPCR | 65 | 169 (150, 192) | 111 (99, 129) |
| GE-E-qPCR | 125 | 124 (110, 140) | 86 (76, 97) |
| MF-E-C-qPCR | 25 | 501 (454, 554) | 333 (301, 372) |
| MF-E-C-qPCR | 45 | 313 (288, 346) | 215 (196, 238) |
| MF-E-C-qPCR | 65 | 244 (224, 270) | 171 (156, 189) |
| MF-E-C-qPCR | 125 | 179 (163, 197) | 129 (118, 143) |
| MF-E-qPCR | 25 | 490 (444, 541) | 328 (292, 361) |
| MF-E-qPCR | 45 | 308 (282, 338) | 210 (192, 231) |
| MF-E-qPCR | 65 | 239 (220, 262) | 167 (154, 184) |
| MF-E-qPCR | 125 | 178 (162, 196) | 129 (117, 141) |
| MF-qPCR | 25 | 472 (429, 523) | 313 (281, 348) |
| MF-qPCR | 45 | 293 (268, 322) | 199 (181, 218) |
| MF-qPCR | 65 | 225 (207, 249) | 156 (143, 173) |
| MF-qPCR | 125 | 164 (149, 181) | 117 (108, 129) |
| MS-E-qPCR | 25 | 352 (316, 395) | 219 (194, 252) |
| MS-E-qPCR | 45 | 212 (193, 238) | 136 (122, 154) |
| MS-E-qPCR | 65 | 162 (146, 182) | 106 (95, 119) |
| MS-E-qPCR | 125 | 118 (105, 135) | 81 (71, 92) |
| TFUF-E-C-qPCR | 25 | 554 (514, 593) | 382 (353, 410) |
| TFUF-E-C-qPCR | 45 | 383 (357, 408) | 273 (256, 292) |
| TFUF-E-C-qPCR | 65 | 315 (293, 338) | 231 (214, 246) |
| TFUF-E-C-qPCR | 125 | 257 (238, 274) | 194 (180, 208) |
| TFUF-E-qPCR | 25 | 559 (516, 602) | 385 (356, 417) |
| TFUF-E-qPCR | 45 | 381 (355, 406) | 272 (253, 289) |
| TFUF-E-qPCR | 65 | 313 (293, 338) | 229 (214, 247) |
| TFUF-E-qPCR | 125 | 255 (234, 273) | 192 (177, 206) |
| TFUF-qPCR | 25 | 542 (501, 585) | 372 (345, 401) |
| TFUF-qPCR | 45 | 370 (344, 396) | 264 (246, 283) |
| TFUF-qPCR | 65 | 305 (283, 327) | 222 (207, 238) |
| TFUF-qPCR | 125 | 243 (226, 264) | 184 (171, 198) |

## Supplement 3: Survey on Collection, Concentration, and Assay Methods for Environmental Surveillance of Salmonella Typhi

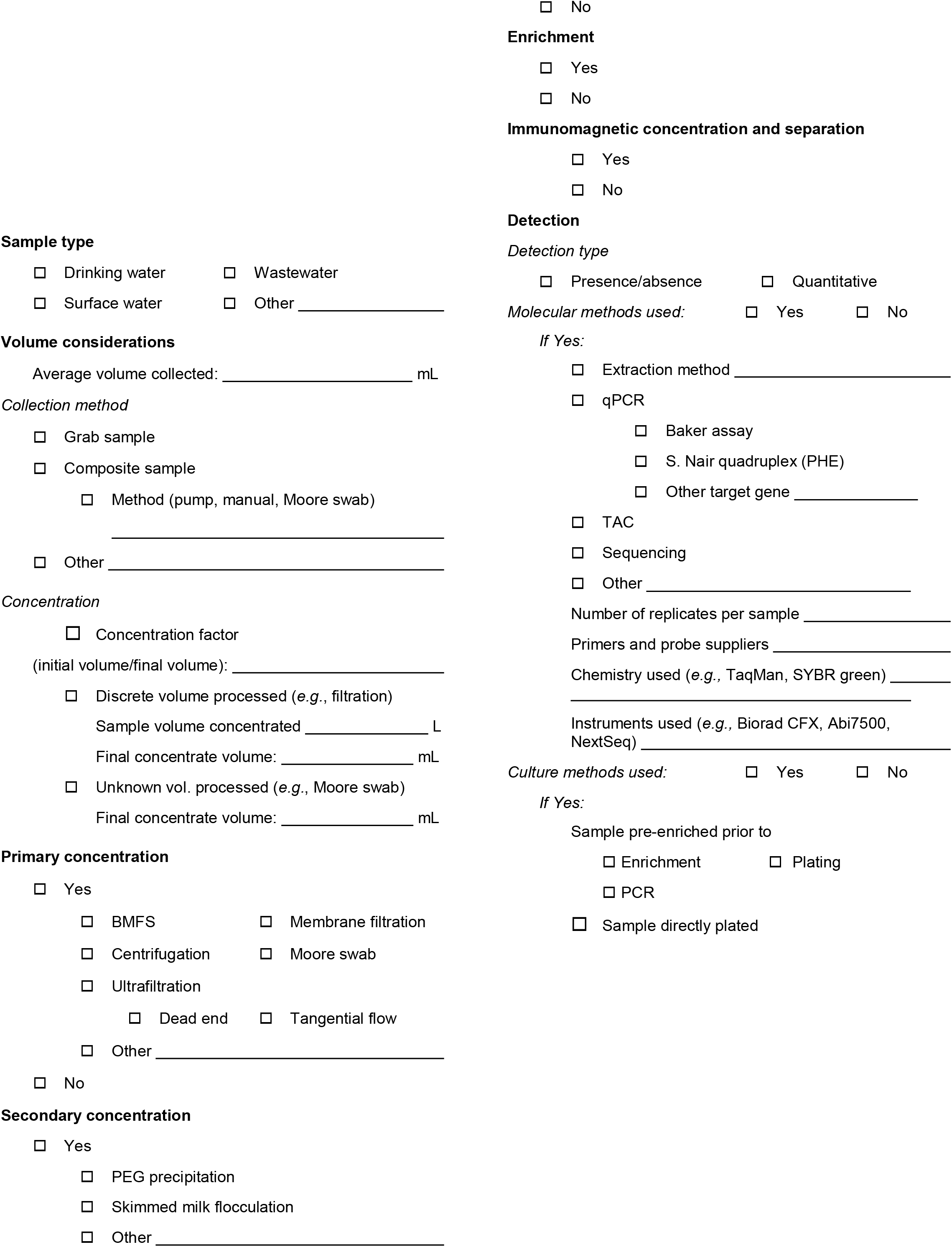
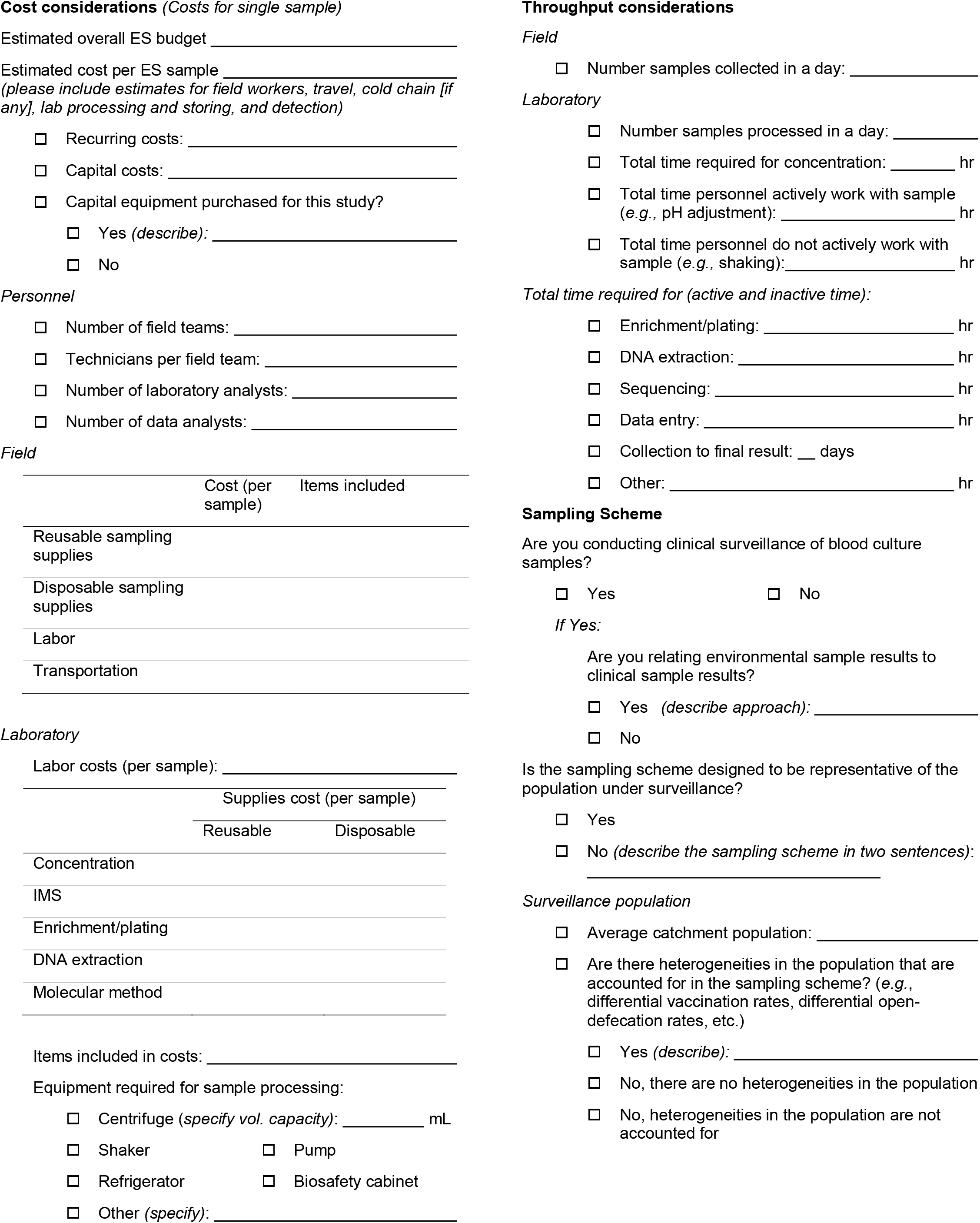

## Supplement 4: Cost Model Details

The total cost model was built per the following formulaic description.

The cost components are structured as follows, where B = binary (zero or one) depending on whether this cost component exists for a given protocol, m = method, M = the number of methods, uc = unit cost, S = the number of samples, and Q = the number of pieces of equipment. Different equipment, labor roles, and consumables are required for different ES methods and only those appropriate for a protocol are costed.

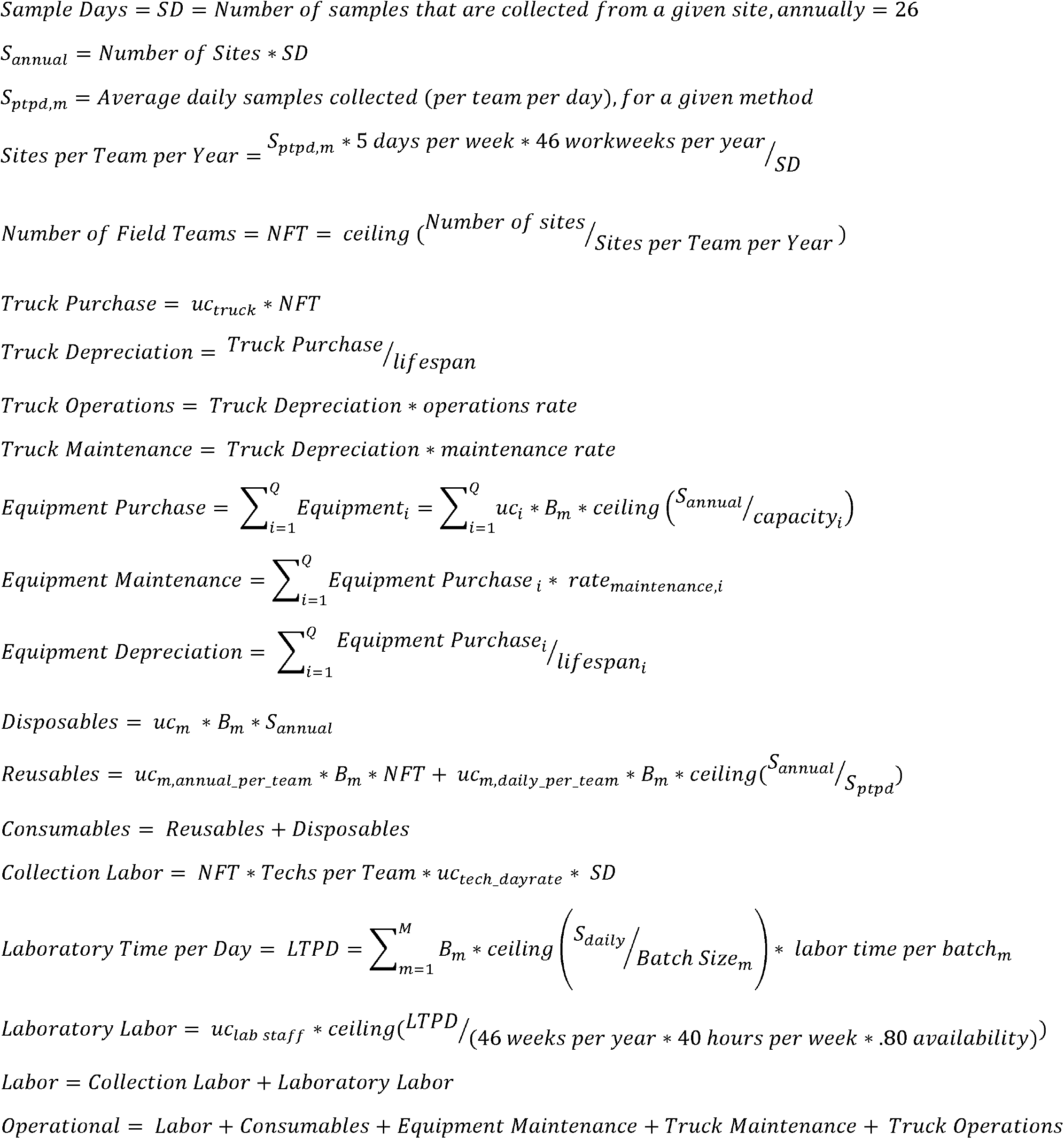
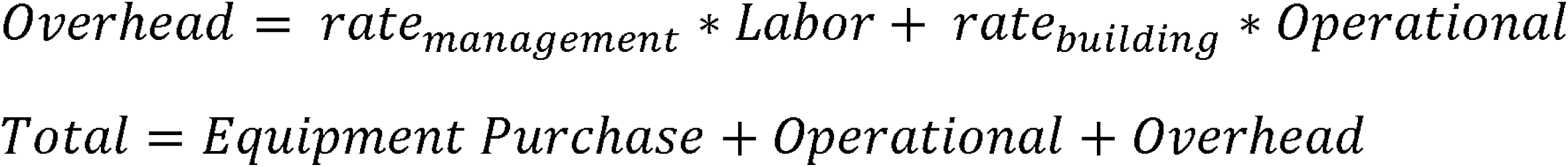

## Notes

### Competing Interest Statement

The authors have declared no competing interest.

### Funding Statement

Dr. Meschke, Dr. Zhou, Ms. Fagnant-Sperati, Mr. Shirai, and Dr. Wang received grant funding from the Bill and Melinda Gates Foundation during the conduct of the study. This publication is based on models and data analysis performed by the Institute for Disease Modeling at the Bill & Melinda Gates Foundation.

